# Separable, symptom specific alterations in brain microstructure associated with early-stage Parkinson’s Disease

**DOI:** 10.1101/2025.07.11.25331216

**Authors:** Mikhail Moshchin, Maureen B Haebig, Cuong P. Luu, Doug C Dean, Alan B. McMillan, Andrew L Alexander, Samuel A Hurley, Catherine L Gallagher, Aaron J. Suminski

**Author notes:** Address for Correspondence: Aaron J. Suminski, 1111 Highland Ave. Rm 3555, Madison, WI 53705. These authors contributed equally to this work (Catherine L Gallagher, Aaron J. Suminski).

## Abstract

**Introduction:** Parkinson’s Disease (PD) is diagnosed based on motor symptoms (bradykinesia, resting tremor, rigidity); yet non-motor symptoms such as sleep abnormalities, autonomic dysfunction, and cognitive changes often precede motor signs, fulfilling the criteria for prodromal PD. How motor and non-motor symptoms emerge from dopamine depletion and whether they involve separable neural substrates remains unclear.

**Methods:** We applied correlational tractography based on multi-shell, diffusion-weighted magnetic resonance imaging in early-stage PD to assess microstructural changes throughout the brain. Eight participants with early-stage PD and 5 healthy controls underwent motor, cognitive, and mood assessments, followed by structural and multi-shell, diffusion-weighted magnetic resonance imaging. Their groupwise differences in white matter integrity associated with PD status were quantified using correlational tractography, with and without age correction.

**Results:** Correlational tractography delineated both microstructural changes that held either a significant positive or negative association with PD status, where the statistical maps of these changes linked differentially to motor and non-motor symptoms. Quantitative anisotropy (QA) extracted from positively associated fibers significantly correlated with cognitive function, while QA of negatively associated fibers correlated with motor function—independent of the effect of age. Of note, QA of positively associated fibers correlated with depressive mood only in the age-uncorrected analyses, suggesting a strong age-related effect.

**Conclusion:** In early-stage PD, motor and non-motor symptoms are mapped to anatomically distinct pathways, suggesting separable pathophysiological mechanisms. These findings further suggest that correlational tractography is appropriate to evaluate changes in structural connectivity in neurodegenerative diseases and, potentially, their therapeutic interventions.

## Introduction

Parkinson’s Disease (PD) is a progressive neurodegenerative disease affecting over 6 million people worldwide. Diagnosed primarily as a movement disorder, PD presents with motor symptoms that canonically include resting tremor, rigidity, and bradykinesia. Motor symptom onset coincides with the degeneration of dopamine-producing neurons in the substantia nigra pars compacta (SNc), resulting in striatal dopamine deficiency [1]. However, recent pathologic studies have revealed that SNc dopamine depletion is preceded by the accumulation of Lewy bodies in the myenteric plexus, olfactory bulb, limbic system, brainstem nuclei, and cerebral cortex [2]. These findings putatively explain the non-motor symptoms that frequently emerge long before the appearance of motor symptoms, including gastrointestinal issues, olfactory dysfunction, sleep disturbances, depression, anxiety, pain, and fatigue [3]. These non-motor symptoms are equally disabling, and they fulfil the criteria for prodromal PD [4]. Thus, there is a critical need to understand the interaction between the motor and non-motor symptoms of PD, especially how they might be instantiated within separable neural substrates.

Our understanding of PD pathophysiology and treatment [5,6] has advanced significantly, thanks to the development of advanced imaging techniques [7]. In particular, imaging reinforced the idea that PD is a disorder of brain networks that profoundly affects brain structure [8,9], function [10], and metabolism [11,12]. For instance, structural studies have shown that, in PD, white matter microstructural integrity loss exceeds that of normal aging and correlates with cognitive symptoms [13,14]. Furthermore, a longitudinal [18F]-fluorodeoxyglucose (FDG) positron emission tomography (PET) study associated mild PD with the activation of two networks with gain and loss of metabolic connections [15]. Along with these human studies, preclinical models corroborate that alterations in a broad brain network are hallmarks of PD (see [16] for review). Our own recent animal work showed that unilateral dopamine depletion leads to microstructural changes in at least two brain networks: one directly related to loss of nigrostriatal dopamine neurons and the other associated with the subsequent compensatory responses [17]. Based on this evidence, we hypothesize that similar network changes could be demonstrated in humans using comparable techniques.

Correlational tractography, a subdivision of connectometry, offers a promising way to evaluate network changes in PD [18]. Unlike techniques assessing end-to-end connections between large parcellated volumes, correlational tractography tracks local connectivity changes to identify fiber tract segments strongly associated with structural, physiological, or behavioral measures. Previous human studies using this approach have focused primarily on motor function within regions-of-interest rather than network-based analyses. When comparing mild PD patients to healthy controls, Wen et al. found significantly lower QA in the corpus callosum, external capsule, cortico-thalamic tracts, and right corticospinal tract [19]. Similarly, Sanchez-Catasus et al. showed impaired integrity of the dopaminergic nigrostriatal system in early-stage PD by combining correlational connectometry and 11C-dihydrotetrabenazine (11C-DTBZ) PET [20]. Kim et al. employed correlational tractography to link basal ganglia, cerebellum, and brainstem changes to motor symptoms, while changes in the temporal and occipital lobes correlated with olfactory dysfunction [21]. As noted, however, research has focused mainly on motor symptoms, with Kim et al. highlighting the need to explore cognitive decline and depression networks in early PD—crucial for developing effective treatments and improving patients’ quality of life.

This project utilized correlational tractography to identify white matter microstructural networks correlated with the disease state in early-stage PD patients and controls. We examined how motor and non-motor symptoms (i.e., cognitive flexibility, attention, and depressive mood) correlated with quantitative anisotropy (QA) within identified networks. Two distinct networks emerged; each linked to either motor symptoms or non-motor symptoms. Notably, these symptom-specific network alterations closely parallel findings from our foundational preclinical study [17], suggesting conserved pathophysiological mechanisms across species and disease models.

## METHODS

### Participants

This pilot study recruited a total of 15 participants, 10 PD patients and 5 healthy controls, through the University of Wisconsin Hospitals and Clinics (Institutional Review Board # 2019- 0987). We selected PD patients with early-stage disease (Hoehn and Yahr stage <= 2) and excluded those with (1) motor symptom onset before age 45; (2) a family history of PD in 2+ first-degree relatives; (3) atypical features including ataxic speech/ limb/ eye signs, supranuclear gaze abnormalities, apraxia, or myoclonus; (4) falls within 2 years of diagnosis; (5) significant cognitive impairment, fluctuating attention, or dementia; or (6) spontaneous hallucinations. For both the PD and control cohort, we further excluded those having (1) an implanted device incompatible with magnetic resonance imaging (MRI), (2) a significant central nervous system disease other than PD (e.g., multiple sclerosis, stroke, brain tumor), or (3) a history of a major psychiatric diagnosis (e.g., schizophrenia, bipolar affective disorder). All participants demonstrated mental capacity before providing informed consent. Data from 2 PD patients were not included in subsequent analyses due to inconsistencies in the acquisition of the imaging data.

### Motor and neuropsychological assessments

All participants underwent motor and cognitive assessments prior to MR imaging. Motor function of PD participants was evaluated while off-medication using the Unified Parkinson’s Disease Rating Scale, Part III Motor Examination (UPDRS III) [22]. The non-motor aspects of PD were assessed while participants were on-medication using 3 tests: the Wisconsin Card Sort Test–64 Card Version to measure cognitive flexibility, working memory, and abstraction (lower score indicates fewer errors) (WCST-64) [23]; the Trail Making Test to evaluate visual attention and task switching (lower score indicates faster time) (TMT) [24]; and the nine-item Patient Health Questionnaire (PHQ-9) to assess the severity of depressive symptoms (lower score indicates lower severity) [25]. Group differences in demographics and assessment scores were evaluated using two-sample t-tests in MATLAB (v2022a).

### MRI acquisition and pre-processing

Following clinical assessments, brain MRIs were captured using a 3T GE SIGNA PET/MR scanner (GE HealthCare, Chicago, IL, USA) with a Nova 32-channel head coil (Nova Medical, Wilmington, MA, USA). In particular, we applied diffusion weighted imaging (DWI) with the following parameters: repetition time (TR) = 6,689 ms, echo time (TE) = 90.3 ms, matrix = 120x120, field of view (FOV) = 24 mm x 24 mm, isotropic voxel size = 2.0x2.0x2.0 mm^3^, multiband acceleration factor R = 3, in-plane acceleration = none, number of excitations (NEX) = 1, three shell acquisition with 9 directions at b=300 s/mm², 18 directions at b=850 s/mm², and 36 directions at b=2,000 s/mm² (Δ = 12.20 ms, δ = 6 ms), with 10 b=0 s/mm^2^ acquisitions interleaved. Scans were repeated twice with reversed phase encoding gradient (AP and PA) to enable correction of susceptibility-induced distortions.

### Image processing and connectometry analysis

DWI data were converted from the DICOM to NIfTI format using dcm2niix [26], then corrected for distortion, eddy current, and motion using FSL TOPUP [27] and EDDY [28,29]. All subsequent analyses were performed using DSI-Studio version Chen-2023-03-18 [18]. We first examined the data for b-table orientation accuracy using an automatic quality control routine [30]. We then reconstructed the data in MNI space using q-space diffeomorphic reconstruction [31] to obtain the spin distribution function [32] with a diffusion sampling of 1.25 and output isotropic resolution of 2.0 mm, employing the ICBM 152 Nonlinear Asymmetrical Template. Subsequently, local connectome matrices were estimated by sampling spin distribution function (SDFs) for each participant using the local fiber directions of the ICBM152 template. Diffusion was quantified using restricted diffusion imaging [33], and quantitative anisotropy (QA) was extracted as a local connectome fingerprint for subsequent correlational tractography analyses [34].

Local connectome matrices (local connectome fingerprints) were assembled into a connectome database and analyzed using two approaches to assess PD-related connectivity changes [35]. In the first, referred to as PD-Age, a non-parametric Spearman correlation evaluated the associations between the local connectome matrices and the PD status. In the second, referred to as PD+Age, a linear regression was first used to remove the effects of participant age on the diffusion data, and then a non-parametric Spearman correlation evaluated the associations between the local connectome matrices and the PD status without contribution from age. T-scores greater than 2.5 were tracked using a deterministic fiber tracking algorithm [36] and filtered with 16 pruning iterations [37]. A false discovery rate (FDR) threshold of 0.05 was used to select tracts significantly associated with the disease state. Tracking used the following parameters: angular threshold = 15 to 90° selected pseudorandomly, step size 0.5 - 1.5 voxel selected pseudorandomly, and tracts shorter than 30 mm or longer than 200 mm were discarded. In total, 1,000,000 seeds were placed. To estimate the false discovery rate (FDR-corrected at p<0.05), a total of 4000 randomized permutations were executed to estimate the null tract length distribution.

Furthermore, we wanted to explore if PD-related fiber tracts show symptom specificity; thus, we examined their association with motor and non-motor symptoms. Correlation tractography previously identified statistically significant and distinct fiber maps associated with the disease state. Now, in each of the PD-Age or PD+Age comparisons for each participant, we extracted the average QA values from fiber tracts that showed a significant association—either positively or negatively—with the disease state. We then fit linear regression models (fitlm in MATLAB) to assess the relationship between the extracted QA values and motor, cognitive, or mood symptoms. Fits were considered significant if an F-test found the slope to be different from zero. Finally, we assessed connectivity patterns by calculating connection strength between regions of interest (ROIs) defined by the FreeSurferSeg Atlas in DSI-Studio. Connection strength was defined as the total number of tracts connecting two ROIs divided by the median length of those tracts. The resulting connectivity matrices from the PD-Age and PD+Age comparisons were superimposed to identify unique versus overlapping fiber tracts. The identity of white matter tracts was assessed using the HCP842 atlas within DSI-Studio.

## RESULTS

### Demographics and cohort characteristics

Of the 15 participants, 13 had imaging data of sufficient quality for analysis (8 PDs and 5 controls; Table 1). PD enrollees were significantly younger than controls (Mean Age: 55.5 vs. 70.4, p=0.006). As expected, PD participants also scored significantly higher on the UPDRS-III compared to controls (Mean score: 15.68 vs. 2.6, p<0.001), reflecting degeneration in their motor function. The WCST-64, TMT(B-A), and PHQ-9 scores showed no significant difference between the PD and control groups.

**Table 1.**
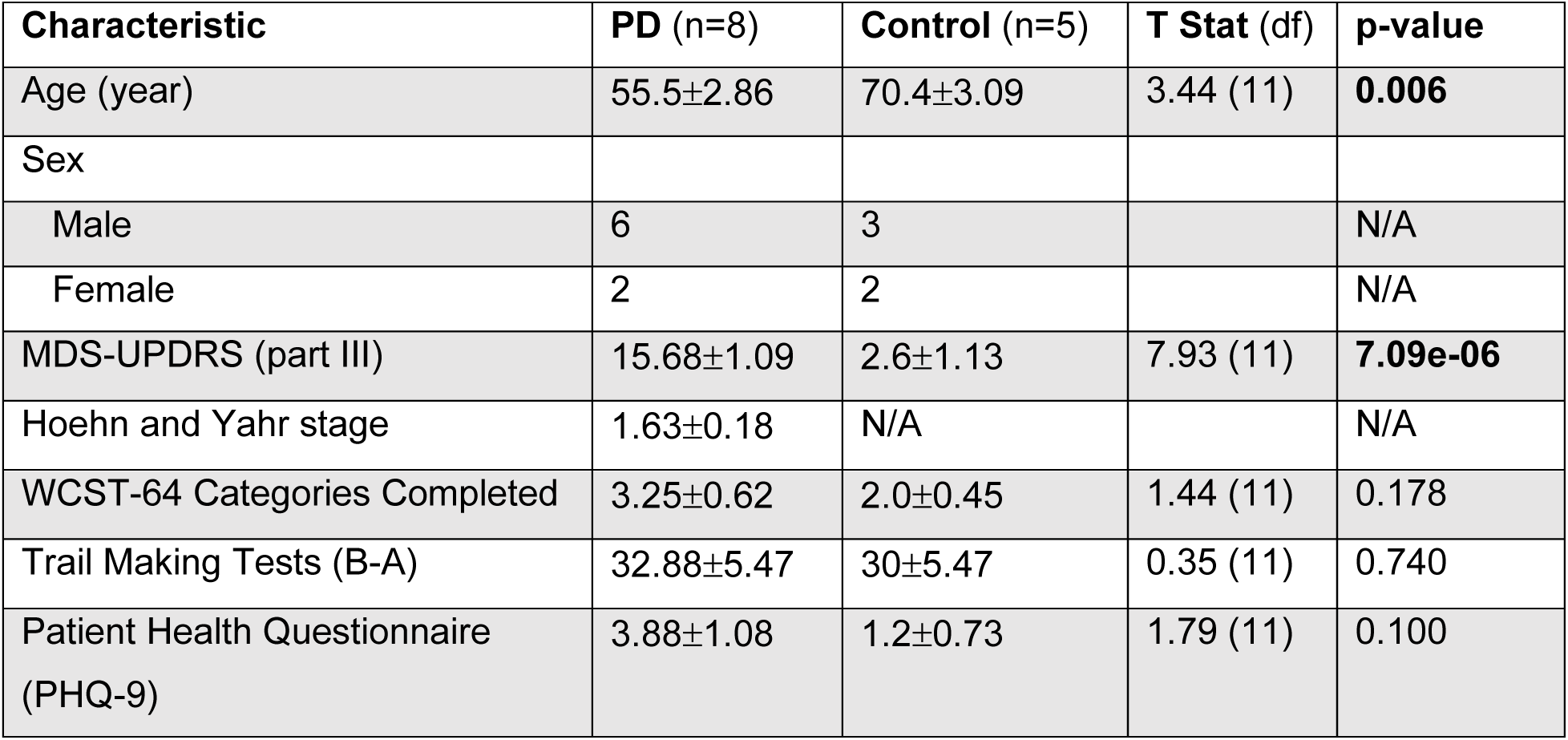
Characteristics of Parkinson’s disease and control participants (Mean±SEM). df = degrees of freedom.

### Parkinson’s Disease modifies structural connectivity throughout the brain

This study aimed to investigate how PD impacts large white matter tracts using correlational tractography. We initially considered the effect of PD on brain microstructure without isolating the effect of participant age (PD-Age; Fig. 1A). Here, correlational tractography showed broad, bilateral networks both positively and negatively associated with PD status. Positively associated fibers (Fig. 1A, Red Fibers) included those in the bilateral prefrontal cortex (BA 10), the forceps minor, corticospinal tract connecting the midbrain, the sensorimotor cortex (BA1-4), and the cerebellar cortex. In contrast, negatively associated fibers (Fig. 1A, Blue Fibers) were most prominent in the midbrain, with strong representation of fibers connecting the basal ganglia and thalamic nuclei. Interestingly, corticospinal tract fibers, distinct from those with a positive association, also showed negative associations with PD status.

**Figure 1.**
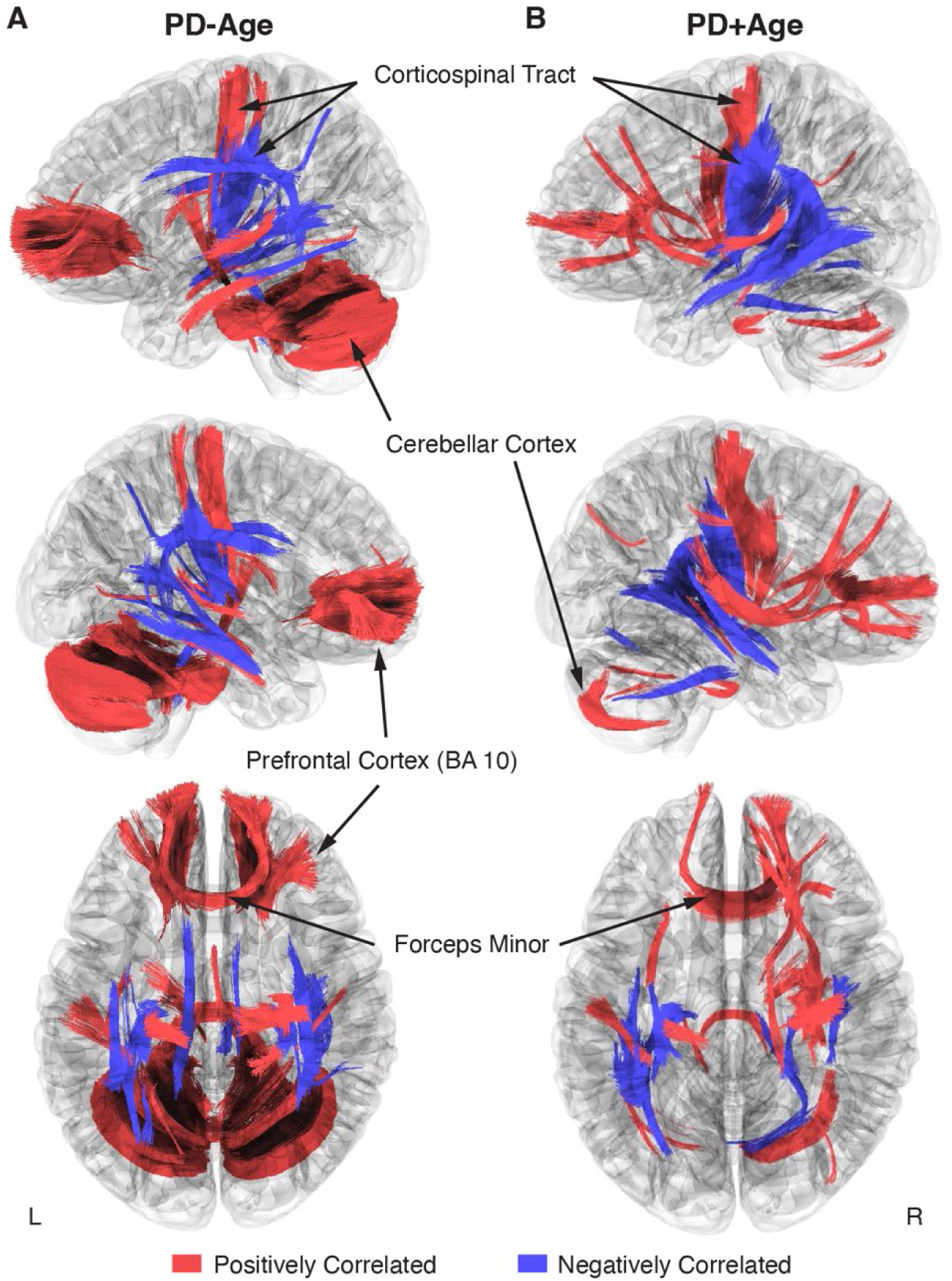
Correlational tractography describing bilateral changes in white matter tracts associated with early-stage PD (FDR<0.05). **A-B**. Overlayed connectomes of positively (red) and negatively (blue) correlated fiber tracts associated with PD status, where **(A)** Age was not isolated as a co-regressor in the PD-Age analysis versus **(B)** Age was controlled in the PD+Age analysis. The first row shows the left hemisphere; the second row shows the right hemisphere.

Because our PD cohort was younger than controls (Table 1), we performed a second correlational tractography analysis controlling for participant age (PD+Age; Fig. 1B). The PD+Age analysis found fewer positively correlated fibers associated with PD status compared to PD-Age (5,138 vs. 44,275 fibers, respectively), while negatively associated fibers remained comparable (3,832 vs. 3,217 fibers, respectively). Importantly, similar network topography was preserved in both analyses, including: positively associated fibers in the bilateral frontal cortex/forceps minor, inferior frontal occipital fasciculus, inferior longitudinal fasciculus, corticospinal tract, and cerebellar peduncle/cortex; as well as negatively associated fibers in the basal ganglia, corticospinal tract, and inferior frontal occipital fasciculus.

When considering the pattern that emerged from the correlational tractography analyses (Fig. 1), positively and negatively associated fibers showed largely distinct anatomical distributions with putatively different functional roles. Positively correlated fibers were predominantly located in higher-order cognitive regions of the prefrontal cortex, cerebellum, and precentral gyrus, whereas negatively correlated fibers were localized in subcortical structures, involving major white matter bundles and projection fibers. Therefore, following our previous preclinical PD work [17], we used linear regression to assess relationships between clinical symptom measures and QA values extracted from these fiber tracts (Fig. 2). In the PD-Age analysis, positively associated fibers showed significant relationships (i.e. regression slopes) with executive function/attention (Trails (B-A), T11 = 2.31, p = 0.04, R^2^=0.33; Fig. 2A, Red Fibers) and depressive mood (PHQ-9, T11 = 2.26, p = 0.045, R^2^=0.32; Fig. 2A, Red Fibers). When controlling for age (PD+Age), the relationship with executive function/attention remained significant (Trails (B-A), T11 = 2.42, p = 0.034, R^2^=0.35; Fig. 2B, Red Fibers) but the depression association was lost (PHQ-9, T11 = 0.84, p = 0.41, R^2^=0.06; Fig. 2B, Red Fibers). We did not observe a significant relationship between the QA of positively correlated fibers and motor function (UPDRS-III) or cognitive flexibility (WCST-64) in either analysis. Interestingly, negatively associated fibers demonstrated significant relationships with motor symptoms in both analyses (PD-Age: UPDRS-III, T11 = -2.67, p = 0.021, R^2^=0.39; Fig. 2A, Blue Fibers) (PD+Age: UPDRS-III, T11 = -2.33, p = 0.039, R^2^=0.33; Fig. 2B, Blue Fibers). The QA of negatively correlated fibers did not show a significant relationship with measures of cognitive flexibility (WCST64), executive function/attention (Trails (B-A)), or depressive mood (PHQ-9) in either analysis.

**Figure 2.**
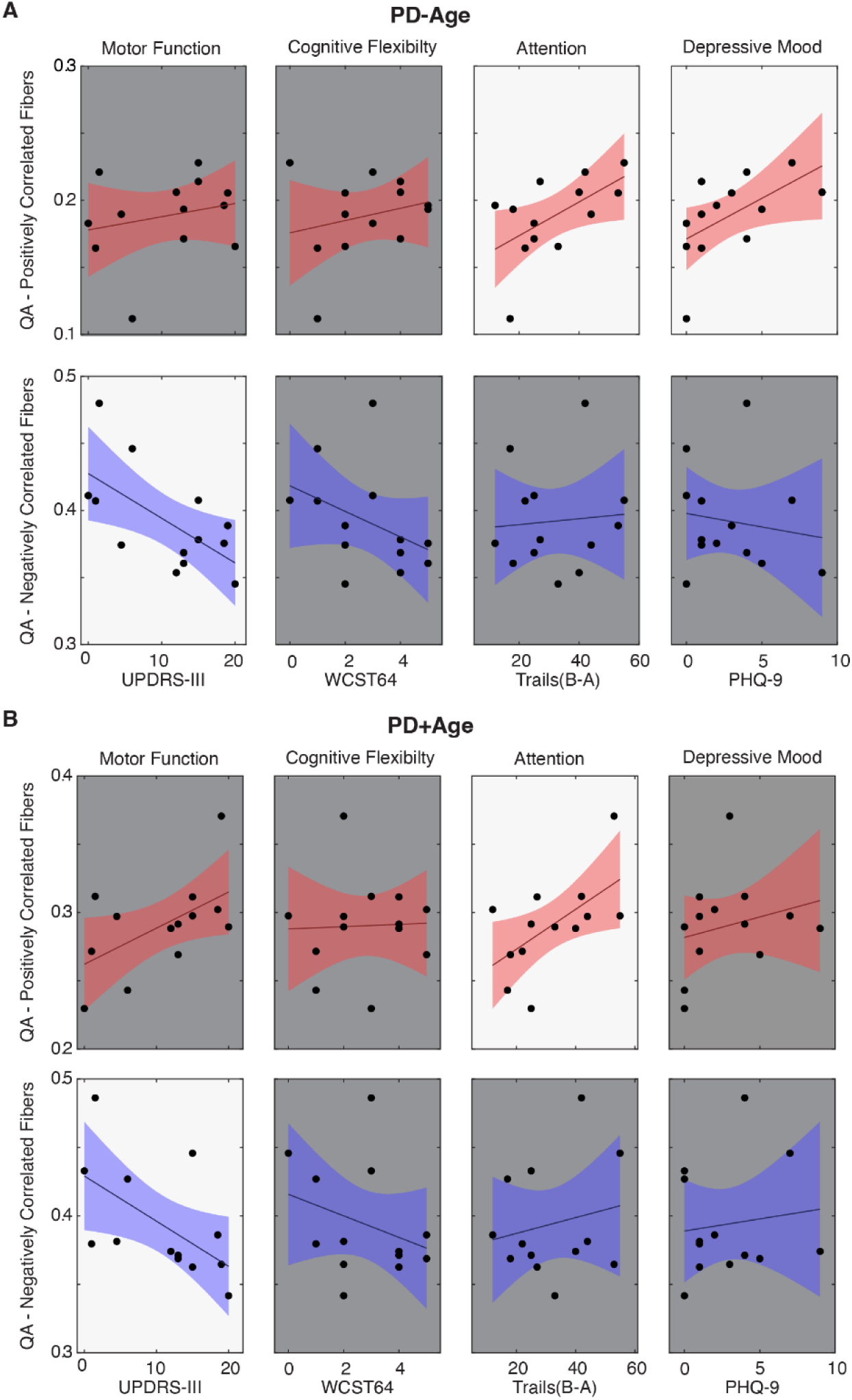
Linear regression models comparing mean QA values of fibers associated with PD status to clinical symptom measures. Panel A-B. UPDRS-III, Wisconsin Card Sort (WST64), Trail Making Tests (B-A), and Patient Health Questionnaire (PHQ-9) were correlated against mean QA of fibers associated with PD status, where the correlational tractography for QA in **(Panel A)** did not isolate Age (PD-Age) and **(Panel B)** controlled for Age (PD+Age). Red indicates fibers associated with increased QA in the early-stage PD cohort, and blue indicates fibers associated with decreased QA (FDR-corrected at p<0.05). A bright background indicates statistical significance (p < 0.05).

Observing that the PD-Age and PD+Age analyses produced statistical maps with different representations of motor and non-motor symptoms (Fig. 2), we then identified brain regions and fiber tracts that were conserved across analyses versus those unique to each analysis. For example, we posited that regions underlying motor disturbances would appear in negatively associated maps of both PD+Age and PD-Age, while depression-related regions would appear solely in the positively correlated map of PD-Age (Fig. 3A). To pursue our goal quantitatively, we extracted separate connectivity matrices for positively and negatively correlated maps from the PD-Age and PD+Age analyses, superimposing them to assess unique different versus overlapping connections (Fig. 3B). We found that motor symptom-related regions were localized to overlapping negatively associated fiber tracts (Fig. 3C, Table S1, Negatively Associated) in the left hemisphere connecting the thalamus, putamen, and pallidum with the primary sensory/motor cortex via the corticospinal tract. These overlapping connections represented a total of 23% (6 of 26) negatively associated connections (Fig. 3C). In contrast, executive/attentional dysfunction in PD was localized to overlapping positively associated fiber tracts (Fig. 3C, Table S1, Positively Associated). Here, these positively associated fibers totaled 35% (39 of 122) of connections and included both intra- and inter-hemispheric connections.

**Figure 3.**
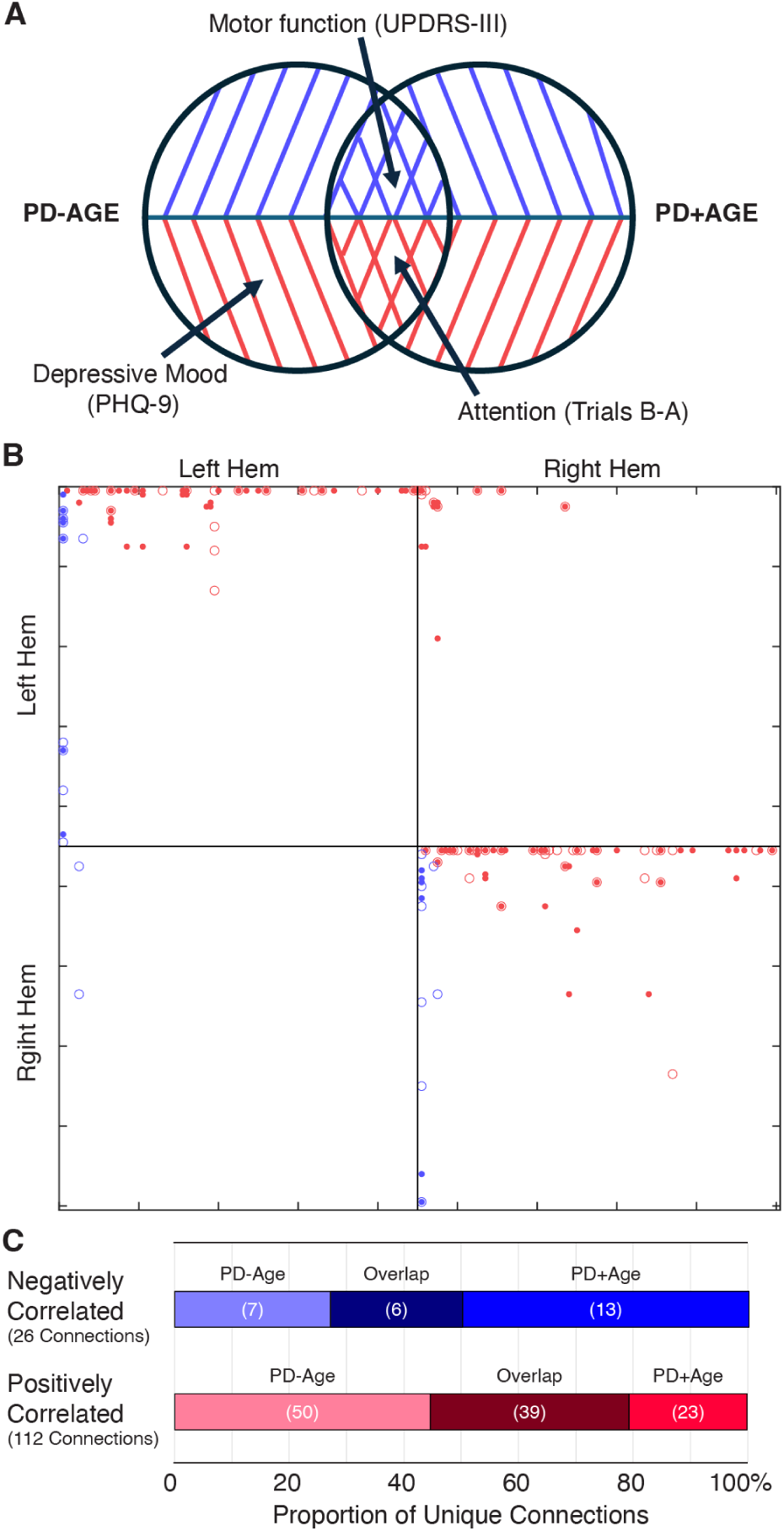
Connection patterns in positively and negatively associated statistical maps support symptom specificity. **A.** Venn diagram illustrating unique versus overlapping connections from the statistical network maps of PD-Age and PD+Age analyses. **B.** Connectivity matrices from correlational tractography using the FreeSurferSeg Atlas in DSI-Studio. Each marker represents tract connections between two regions of interest (ROIs), where: red indicates positively associated versus blue indicating negatively associated fiber tracts; dots represent PD-Age versus open circles representing PD+Age analysis results (age-controlled). **C.** All unique connections (from each of the positively or negatively correlated groups) were further categorized based on whether they appeared solely in the PD-Age, solely in the PD+Age, or in both analyses (Overlap).

Intrahemispheric connections included the right cerebellar cortex, right corticospinal tract linking subcortical structures to the primary motor cortex, a smaller portion of the left corticospinal tract innervating the primary motor cortex, and the right prefronto-caudate tract extending to the middle frontal and orbital gyrus. Overlapping interhemisphic connections were limited to forceps minor connecting the superior frontal gyrus and transverse frontopolar gyrus/sulcus. Because depressive mood was only related to positively associated fibers in the PD-Age analysis, we focused on the unique connections in the PD-Age statistical map to localize brain regions and fiber tracts that may directly relate to depressive mood (Fig. 3B). The unique connections positively associated with PD-Age status (Fig. 3A, Table S2) included interhemispheric connections between (1) pons and bilateral cerebellar cortex via middle cerebellar peduncle, and (2) middle frontal gyrus/sulcus via forceps minor and accounted for 45% (50 of 119) of all connections.

## Discussion

While PD is diagnosed based on motor dysfunction, early prodromal symptoms are nonspecific, manifesting as cognitive, olfactory, and gastrointestinal changes. Furthermore, the pathophysiology of these non-motor symptoms and their associated brain network changes remains less understood than that of motor symptoms. This study aimed to identify disease-correlated structural networks using diffusion-based correlational tractography during early-stage PD and examine how QA within these networks correlated with motor and non-motor symptoms.

Correlational tractography demonstrated separable statistical maps positively and negatively associated with PD status (Fig. 1). QA within these networks showed differential relationships with motor and non-motor symptom severity, measured using standardized clinical scales.

Furthermore, cognitive dysfunction and depressive mood were associated with a specific network connecting the prefrontal cortex (BA10) bilaterally, the right prefrontal cortex (BA8), the basal ganglia, and the cerebellum. In contrast, motor dysfunction, measured by UPDRS-III, was correlated with regions/ tracts showing negative association with the disease state, specifically linking the basal ganglia with the sensorimotor cortex via the corticospinal tract. Our findings suggest that structural white matter dysregulation/ damage observed on correlational tractography may contribute to both non-motor (i.e., cognitive/ depressive) and motor differences detected in early-stage PD patients.

### Symptom-specific brain networks in PD

Numerous studies have reported associations between functional/ structural alterations and clinical measures of both motor and non-motor symptoms [13,14,19,20,38,39]. For example, a recent longitudinal positron emission tomography (PET) study by Tang et al. illustrated the activation of two distinct brain networks, each characterized by either gain or loss of metabolic connections in early-stage PD [40]. Consistent with Tang et. al., our analysis in early-stage PD patients revealed two distinct statistical brain maps, one correlated with motor symptoms and the second with non-motor symptoms. Likewise, Kim, et. al. showed that variations in QA within separable brain regions were selectively correlated with either motor or non-motor (i.e. olfactory) symptoms of PD. Notably, both age-controlled (PD+Age) and uncorrected (PD-Age) correlational tractography analyses revealed robust associations for motor and non-motor functions in similar brain regions, suggesting that these symptom-specific networks are independent of age effects.

#### Motor symptoms

We demonstrated that overlapping statistical network maps of motor symptoms in PD, maps common in both the PD-Age and PD+Age analyses, were linked to negatively associated fiber tracts in the left hemisphere (Table S1, Negatively Associated). These tracts connect the thalamus, putamen, and pallidum with the primary sensory and motor cortex via the corticospinal tract, all of which play critical roles in motor control [41] and show well-recognized dysfunction in PD (see [42] for review). Our findings align with previous studies identifying motor-related structural changes. Interestingly, Kim et al. found that QA from the basal ganglia, brainstem, thalamus, and cerebellum most strongly correlated with motor symptoms (UPDRS-III) in early-stage PD; however, their analysis did not distinguish between individual nuclei, limiting direct comparison. Similar to our study, Sanchez-Catasus et al. used correlational tractography to identify nigrostriatal fibers associated with striatal dopamine denervation measured by ^11^C-dihydrotetrabenazine PET [20]. They showed that QA extracted from these fibers was negatively correlated with bradykinesia severity in patients with mild to moderate PD, suggesting that the reduced QA seen with increased disease severity reflects impaired axonal integrity in the nigrostriatal pathway. Our recent preclinical work corroborates the correlation between reduced QA and impaired integrity of the nigrostriatal dopamine pathway [17,43].

Following unilateral 6-hydroxydopamine lesions in the median forebrain bundle (MFB), we showed that QA from the MFB strongly correlated with the degeneration of dopaminergic projections from the SNc to the striatum, measured via tyrosine hydroxylase staining. Taken together, these results demonstrate that impaired motor function in PD patients is strongly associated with degeneration of white matter tracts connecting the midbrain to the sensorimotor cortical areas. Future preclinical and clinical research should focus on the ability of this methodology to ascertain the efficacy of medical or neuromodulation therapy with the goal of optimizing patient specific interventions.

#### Non-motor symptoms

Similar to motor symptoms, we explored whether the statistical maps identified using correlational tractography carry significant correlations with non-motor symptoms of PD, including cognitive flexibility, attention, and depressive mood. We found no relationship between QA extracted from both positively and negatively associated maps with the WCST64, a measure of executive function. This is not surprising given that the decline on the WCST64 is most pronounced in late-stage PD [44], while our study cohort included only patients in early-stage PD.

In contrast, decreased attention/working memory function, measured by TMT, strongly correlated with QA extracted from fibers positively associated with PD status in both the PD-Age and PD+Age analyses. Though group-wise comparison of our two study cohorts did not show a significant difference in TMT (B-A), previous studies support attention/ working memory dysfunction in PD patients [45]. Hanganu et al. found that PD patients with cognitive impairment exhibited increased diffusivity metrics in fiber bundles connecting the dorsolateral prefrontal cortex to subcortical structures, including the thalamus, caudate, and putamen [46]. Similarly, using metabolic and structural studies in PD patients, Kubler et al. showed significant degeneration of frontostriatal networks and reduced metabolism in the prefrontal cortex, with TMT performance correlating with measures of prefrontal diffusion. In both PD+Age and PD- Age analyses, regions associated with deficits in attentional and executive control were most prominent within fiber tracts positively associated with PD (Table S1, Positively Associated).

Importantly, these included connections via the forceps minor that link the bilateral prefrontal cortex, specifically the superior frontal gyrus, the transverse frontopolar gyrus/sulcus, and the right prefronto-caudate tract. Indeed, Dirnberger et al. have put forth a plausible interpretation: dysfunction in the basal ganglia of PD patients, similar to our negatively correlated networks, necessitates compensatory recruitment of executive regions for tasks healthy individuals perform automatically—where the recruited fibers are represented by our ‘overlapping’ positively associated fibers [47].

Depression is strongly linked to PD [19,48,49]. For example, a recent meta-analysis of >38,000 subjects showed that one in three PD patients were diagnosed with depression. Multifactorial in origin, depression in PD seems to be strongly related to symptom severity and age of onset [49], more common with disabling axial motor symptoms like postural instability and gait disorders [48]. Mood disorders have been linked to functional changes in limbic-cortical function, particularly in the subgenual cingulate and prefrontal cortex [50]. Ansari et al. proposed that early depressive symptoms in PD are reactive responses to chronic degeneration, suggesting that the prefrontal and orbitofrontal cortices may be involved in early PD depression [51].

Furthermore, Prange et al. reported that depressive symptoms in early-stage PD patients are associated with altered diffusivity in the right limbic subnetwork, including the genu of the corpus callosum and the forceps minor [52]. Importantly, these are the regions most targeted for deep brain stimulation in treatment-resistant depression [53,54]. Our data highlighted the correlation between depressive symptoms and the connections between (1) the middle frontal gyrus/sulcus via the forceps minor and (2) the pons and bilateral cerebellar cortex via the middle cerebellar peduncle (Figs. 2 and 3). Importantly, these connections were unique to PD-Age analysis and absent when controlling for age (PD+Age), aligning with past findings and reinforcing an age-dependent depression mechanism in early PD. The alignment with past findings regarding age further reinforces the validity of correlational tractography in detecting early-stage PD.

## Limitations

Using correlational tractography, we demonstrated that structural changes in early-stage PD include two distinct networks associated with motor versus non-motor symptoms. Several key limitations need to be addressed in this study. First, this is an early-stage pilot exploratory study aiming to assess whether correlational tractography based on multi-shell dMRI can identify symptom-specific brain networks in PD. As a result, the regression analyses used comparisons uncorrected for multiple comparisons. Future confirmatory studies with a larger sample size are necessary to confirm the results of this work. That said, this report is timely given recent preclinical work showing similar symptom-specific networks in the unilateral 6-OHDA animal model [17]. Second, our control cohort was significantly older than the PD cohort, potentially confounding results due to age-related brain changes. Our analysis of both age-corrected and uncorrected networks, however, helped distinguish age-dependent from age-independent PD effects, ultimately finding many similar fiber maps. Finally, we used the ICBM152 template derived from young adults; future studies should employ customized demographic-matched templates to improve accuracy and generalizability.

## Data Availability

All data produced in the present study are available upon reasonable request to the authors.

## Acknowledgement

Support for this research was provided by the Departments of Radiology, Neurology and Neurological Surgery at the University of Wisconsin-Madison.

The authors have no conflicts to declare.

