## Supplementary Tables for "Separable, symptom specific alterations in brain microstructure associated with early-stage Parkinson’s Disease"

**Table S1.** Overlapping connections between PD-Age and PD+Age correlational tractography analyses reveal putative symptom-specific networks for motor and cognitive impairments in early-stage PD.

**Common Connections in Positively Associated Statistical Maps**

| Hemisphere | Area | Hemisphere | Area |
|---|---|---|---|
| L | Anterior transverse collateral sulcus | L | Cerebral White Matter |
| L | Central sulcus (Rolando's fissure) | L | Cerebral White Matter |
| R | Central sulcus (Rolando's fissure) | R | Cerebral White Matter |
| R | Central sulcus (Rolando's fissure) | R | Pallidum |
| R | Fronto-marginal gyrus (of Wernicke) and sulcus | R | Cerebral White Matter |
| R | Lateral occipito-temporal gyrus | L | Cerebellum-Cortex |
| R | Lateral occipito-temporal gyrus | R | Cerebellum-Cortex |
| R | Middle frontal gyrus (F2) | R | Cerebral White Matter |
| R | Orbital gyri | R | Cerebral White Matter |
| R | Orbital part of the inferior frontal gyrus | R | Cerebral White Matter |
| L | Paracentral lobule and sulcus | L | Cerebral White Matter |
| L | Temporal plane of the superior temporal gyrus | L | Cerebral White Matter |
| L | Precentral gyrus | L | Cerebral White Matter |
| R | Precentral gyrus | R | Cerebral White Matter |
| R | Precentral gyrus | R | Pallidum |
| L | Superior frontal gyrus (F1) | L | Cerebral White Matter |
| R | Superior frontal gyrus (F1) | R | Cerebral White Matter |
| L | Superior temporal sulcus (parallel sulcus) | L | Cerebral White Matter |
| R | Superior temporal sulcus (parallel sulcus) | R | Cerebral White Matter |
| R | Transverse frontopolar gyri and sulci | L | Cerebral White Matter |
| R | Transverse frontopolar gyri and sulci | R | Cerebral White Matter |
| R | Transverse frontopolar gyri and sulci | R | Choroid-plexus |
| L | Transverse temporal sulcus | L | Cerebral White Matter |
| L | Thalamus-Proper | L | Cerebral White Matter |
| L | Putamen | L | Cerebral White Matter |
| L | Pallidum | L | Cerebral White Matter |
| L | VentralDC | L | Cerebral White Matter |
| L | VentralDC | L | Thalamus-Proper |
| R | Cerebral-White-Matter | L | Cerebral White Matter |
| R | Lateral-Ventricle | R | Cerebral White Matter |
| R | Cerebellum-White-Matter | L | Cerebellum-White-Matter |
| R | Cerebellum-Cortex | L | Cerebellum-Cortex |
| R | Cerebellum-Cortex | R | Cerebellum-White-Matter |
| R | Thalamus-Proper | R | Cerebral White Matter |
| R | Putamen | R | Cerebral White Matter |
| R | Pallidum | R | Cerebral White Matter |
| R | VentralDC | R | Cerebral White Matter |
| R | Choroid-plexus | L | Cerebral White Matter |
| R | Choroid-plexus | R | Cerebral White Matter |

**Common Connections in Negatively Associated Statistical Maps**

| Hemisphere | Area | Hemisphere | Area |
|---|---|---|---|
| L | Cerebral White Matter | L | Thalamus-Proper |
| L | Cerebral White Matter | L | Putamen |
| L | Cerebral White Matter | L | Pallidum |
| L | Cerebral White Matter | L | VentralDC |
| L | Cerebral White Matter | L | Anterior segment of the circular sulcus of the insula |
| R | Cerebral White Matter | R | Superior temporal sulcus (parallel sulcus) |

**Table S2.** Unique connections to the PD-Age correlational tractography analyses reveal putative symptom-specific networks for depressive mood symptoms.

| Hemisphere | Area | Hemisphere | Area |
|---|---|---|---|
| R | Anterior part of the cingulate gyrus and sulcus | R | Cerebral White Matter |
| R | Anterior transverse collateral sulcus | R | Cerebral White Matter |
| L | Fronto-marginal gyrus (of Wernicke) and sulcus | L | Cerebral White Matter |
| L | Fronto-marginal gyrus (of Wernicke) and sulcus | L | Choroid-plexus |
| R | Fronto-marginal gyrus (of Wernicke) and sulcus | R | Caudate |
| R | Fronto-marginal gyrus (of Wernicke) and sulcus | R | Putamen |
| L | Lateral aspect of the superior temporal gyrus | L | Cerebral White Matter |
| R | Lateral aspect of the superior temporal gyrus | R | Cerebral White Matter |
| L | Lateral occipito-temporal gyrus | L | Cerebellum-Cortex |
| R | Lateral orbital sulcus | R | Cerebral White Matter |
| L | Lingual gyrus/medial occipito-temporal gyrus | L | Cerebellum-White-Matter |
| L | Lingual gyrus/medial occipito-temporal gyrus | L | Cerebellum-Cortex |
| R | Lingual gyrus/medial occipito-temporal gyrus | R | Cerebellum-Cortex |
| R | Lingual gyrus/medial occipito-temporal gyrus | R | Lateral occipito-temporal gyrus |
| L | Middle frontal gyrus (F2) | L | Cerebral White Matter |
| L | Middle frontal gyrus (F2) | L | Lateral-Ventricle |
| L | Middle frontal sulcus | L | Cerebral White Matter |
| R | Middle frontal sulcus | R | Cerebral White Matter |
| R | Occipital pole | R | Lateral occipito-temporal gyrus |
| L | Orbital gyri | L | Cerebral White Matter |
| R | Orbital gyri | R | Transverse frontopolar gyri and sulci |
| L | Orbital sulci (H-shaped sulci) | L | Cerebral White Matter |
| R | Orbital sulci (H-shaped sulci) | R | Cerebral White Matter |
| R | Orbital sulci (H-shaped sulci) | R | Putamen |
| R | Paracentral lobule and sulcus | R | Cerebral White Matter |
| R | Pericallosal sulcus (S of corpus callosum) | R | Cerebral White Matter |
| R | Postcentral gyrus | R | Cerebral White Matter |
| L | Straight gyrus, Gyrus rectus | L | Cerebral White Matter |
| L | Suborbital sulcus | L | Cerebral White Matter |
| R | Suborbital sulcus | R | Cerebral White Matter |
| L | Subparietal sulcus | L | Cerebral White Matter |
| L | Superior frontal gyrus (F1) | L | Lateral-Ventricle |
| L | Superior frontal gyrus (F1) | L | Choroid-plexus |
| R | Superior frontal gyrus (F1) | R | Choroid-plexus |
| L | Transverse frontopolar gyri and sulci | L | Cerebral White Matter |
| L | Transverse frontopolar gyri and sulci | L | Lateral-Ventricle |
| L | Transverse frontopolar gyri and sulci | L | Choroid-plexus |
| L | Lateral-Ventricle | L | Cerebral White Matter |
| L | Cerebellum-Cortex | L | Cerebellum-White-Matter |
| L | Caudate | L | Cerebral White Matter |
| L | VentralDC | L | Putamen |
| L | VentralDC | L | Pallidum |
| L | Choroid-plexus | L | Cerebral White Matter |
| R | Cerebral White Matter | L | Choroid-plexus |
| R | Lateral-Ventricle | L | Choroid-plexus |
| R | Cerebellum-White-Matter | L | Cerebellum-Cortex |
| R | Cerebellum-Cortex | L | Cerebellum-White-Matter |
| R | Cerebellum-Cortex | L | Lingual gyrus/medial occipito-temporal gyrus |
| R | Caudate | R | Cerebral White Matter |
| R | Choroid-plexus | R | Lateral-Ventricle |
